# Examining face-mask usage as an effective strategy to control COVID-19 spread

**DOI:** 10.1101/2020.08.12.20173047

**Authors:** Adam Catching, Sara Capponi, Ming Te Yeh, Simone Bianco, Raul Andino

**Author notes:** authors contributed equally to the manuscript (Adam Catching, Sara Capponi).

## Abstract

COVID-19’s high virus transmission rates have caused a pandemic that is exacerbated by the high rates of asymptomatic and presymptomatic infections. These factors suggest that face masks and social distance could be paramount in containing the pandemic. We examined the efficacy of each measure and the combination of both measures using an agent-based model within a closed space that approximated real-life interactions. By explicitly considering different fractions of asymptomatic individuals, as well as a realistic hypothesis of face masks protection during inhaling and exhaling, our simulations demonstrate that a synergistic use of face masks and social distancing is the most effective intervention to curb the infection spread. To control the pandemic, we show that practicing social distance is less efficacious than the widespread usage of face masks and that wearing face masks provides optimal protection even if only a small portion of the population comply with social distance. Finally, the face mask effectiveness in curbing the viral spread is not reduced if a large fraction of population is asymptomatic. Our findings have important implications for policies that dictate the reopening of social gatherings.

**Author summary:** The COVID-19 outbreak has created an enormous burden on the worldwide population. Among the various ways of preventing the spread of the virus, face masks have been proposed as a main way of reducing transmission. Yet, the interplay between the usage of face mask and other forms of Non-Pharmaceutical Intervention is still not completely clear. In this paper we introduce a stochastic individual-based model which aims at producing realistic scenarios of disease spread when mask wearing with different inward and outward efficacy and social distancing are enforced. The model elucidates the conditions that makes the two forms of intervention synergistic in preventing the spread of the disease.

## Introduction

The COVID-19 outbreak has caused high levels of mortality and economic damage around the world. The causative agent of COVID-19, SARS-CoV-2, is an airborne pathogen that can be transmitted between humans through droplets and aerosols that can travel 1–8 meters^1^. The virus is transmitted by both symptomatic and asymptomatic individuals. COVID-19 can cause severe symptoms that require hospitalization in 1-5% of cases, as well as severe long-term sequels and death. Accordingly, the outbreak has seriously impacted healthcare systems around the world^2^. One of the major difficulties to contain the COVID-19 pandemic has been detection of infected asymptomatic or pre-symptomatic individuals, who are estimated to be responsible for as much as 95% of all transmissions^34^. As these individuals carry and spread the virus without manifesting any sign of the disease, they represent a crucial variable in managing the outbreak.

In the absence of an effective vaccine or antiviral, most countries have implemented non-pharmaceutical interventions (NPIs) to curb the spread of COVID-19^5^. These include closure of schools, workplaces, churches, offices, factories and other social venues, while encouraging preventative measures ranging from maintaining social distancing (SD, usually 2 m/6 ft between individuals) to total quarantine and societal lockdowns. These measures aim to reduce the effective contact rate of the population, which in turn decreases the disease reproductive number R_e_. These NPIs limit the epidemic, but they present important drawbacks. Total lockdown can only be implemented for short periods, due to its severe impact on the social fabric and economy of a country. Meanwhile, essential workers remain vulnerable to infection and transmission due to the frequent encounters with infected, often asymptomatic, individuals. Because of their social and economic impact, lockdowns and SD measures have been lifted in some countries, leading to reactivation of virus spread and ensuing increased morbidity and mortality.

Face masks covering the nose and mouth area also provide a level of filtration that blocks virus transmission to a certain extent^6-8^. Masks prevent the spread of droplets and aerosols generated by an infected individual^1^, where correctly worn surgical masks can reduce viral transmission by 95%. Uninfected individuals wearing a surgical mask are about 85% protected against infection^9^. Masks may be more effective than restrictions in people’s interactions for controlling the spread of infectious virus because they prevent the larger expelled droplets from being converted into smaller droplets that can travel farther, rather than removing the interactions between individuals that cause droplets. Accordingly, face masks reduce the spread of influenza^10^ and coronaviruses^11,12^.

In the past, several papers have used theoretical models to study how efficacious mask wearing is in avoiding the spread of airborne viruses. In 2010 at least two studies were focused on the effect of face mask usage to contrast the diffusion of Novel Influenza A (H1N1)^13,14^. Although these works are not on COVID-19 or another Coronavirus disease, they underline the necessity of developing more accurate models to describe similar diseases and reinforce their results in the light of new diseases. Specifically on COVID-19, the efficacy of mask wearing has recently been studied using an ordinary differential equations (ODE) model^15^, which considered also a varying percentage of asymptomatic individuals, compliance with mandate to wear masks, and a different inward and outward efficacy of protection. However, the authors consider face masks as the sole preventative, excluding additional NPIs like SD or shelter in place^1^.

Additionally, the problem has been studied in several papers using agent-based models, each being focused on a specific part of the problem. In some cases, the presence of asymptomatic infected individuals was not considered^16,17^. In other studies, the efficacy of wearing masks was not analyzed when combined with other NPIs^18^. Finally, the difference in inward and outward protection given by a face mask was often neglected or parametrized with a single value^19^. A rather comprehensive data-driven investigation of all these effects has been performed by Hoertel et al.^20^. However, their results are specific to the country of France, and the high dimensionality of the parameter space makes it difficult to disentangle the effect of the various interventions.

In our current study, we analyzed the relative efficacy of wearing face masks and/or exercising SD to reducing the spread of COVID-19 in the presence of asymptomatic individuals. This analysis may be particularly important in the current phase of the pandemic. As pharmaceutical interventions (e.g. vaccines) are deployed, people may experience a false sense of safety, which may lead to unsafe behavior. This may allow the virus to circulate at the interface between immune and non-immune individuals, accelerating the emergence of variants resistant to vaccination. Face masks have distinct inward and outward protection, parameterized using a Gamma function (see Methods). Through stochastic computer simulations of infection spread, we modeled realistic outbreak scenarios and found that SD only yields beneficial effects if accompanied by a widespread population adherence to SD. In contrast, wearing face masks is a highly effective strategy to reduce the spread of infection. Our results are general, and suggest that, even when a large fraction of infected individuals is asymptomatic, mask wearing is the most effective strategy to control virus spread and alleviate the impact of COVID-19 outbreak, particularly when combined with conditions of partial SD compatible with the function of society.

## Results

### Stochastic model description and calibration

We developed an agent-based model (ABM) to examine the effectiveness of wearing masks and SD on the rate of infections and viral spread during the pandemic. Unlike ODE models of the spread of COVID-19 disease^15,21^, ABMs are stochastic models that allow the description of non-homogeneous distributions of agents that act individually^12,22^. In ABMs, each individual behaves dynamically and independently in response to environmental changes^23^ according to rules that describe their interactions. To model the COVID-19 pandemic, we used a SEAIR system in which each agent represents an individual who can be susceptible (S), exposed (E), asymptomatically infected (A), symptomatically infected (I), or recovered (R). Although important, we do not explicitly consider reinfection with the same or a different viral variant, as it is outside the scope of this work. To obtain a realistic model of virus spread, we chose parameters that describe the spread of SARS-CoV2: transmission events occur through contacts made between susceptible and infectious individuals in close proximity (distance ≤ 2*r*, Fig. 1A), and exposed individuals undergo an incubation period of 5.1 days to become infectious (Fig. 1, spheres with red border). This incubation time represents the interval required to increase viral loads to levels sufficient for transmission^34,24^. We assume that recovered individuals (Fig. 1, spheres with black border), who resolve the infection, cannot be re-infected or infect others, which is a reasonable assumption for the duration of our models (45 days). Symptomatic agents are in the Infected state for 7 days but are only infectious to other agents for 12 hours. Asymptomatic agents are infectious for all 7 days but the probability of transmission is reduced by 33%. Presymptomatic agents are similar to Susceptible agents in their interactions but have a countdown for 5.1 days until they become either asymptomatically or symptomatically Infected

**Figure 1.**
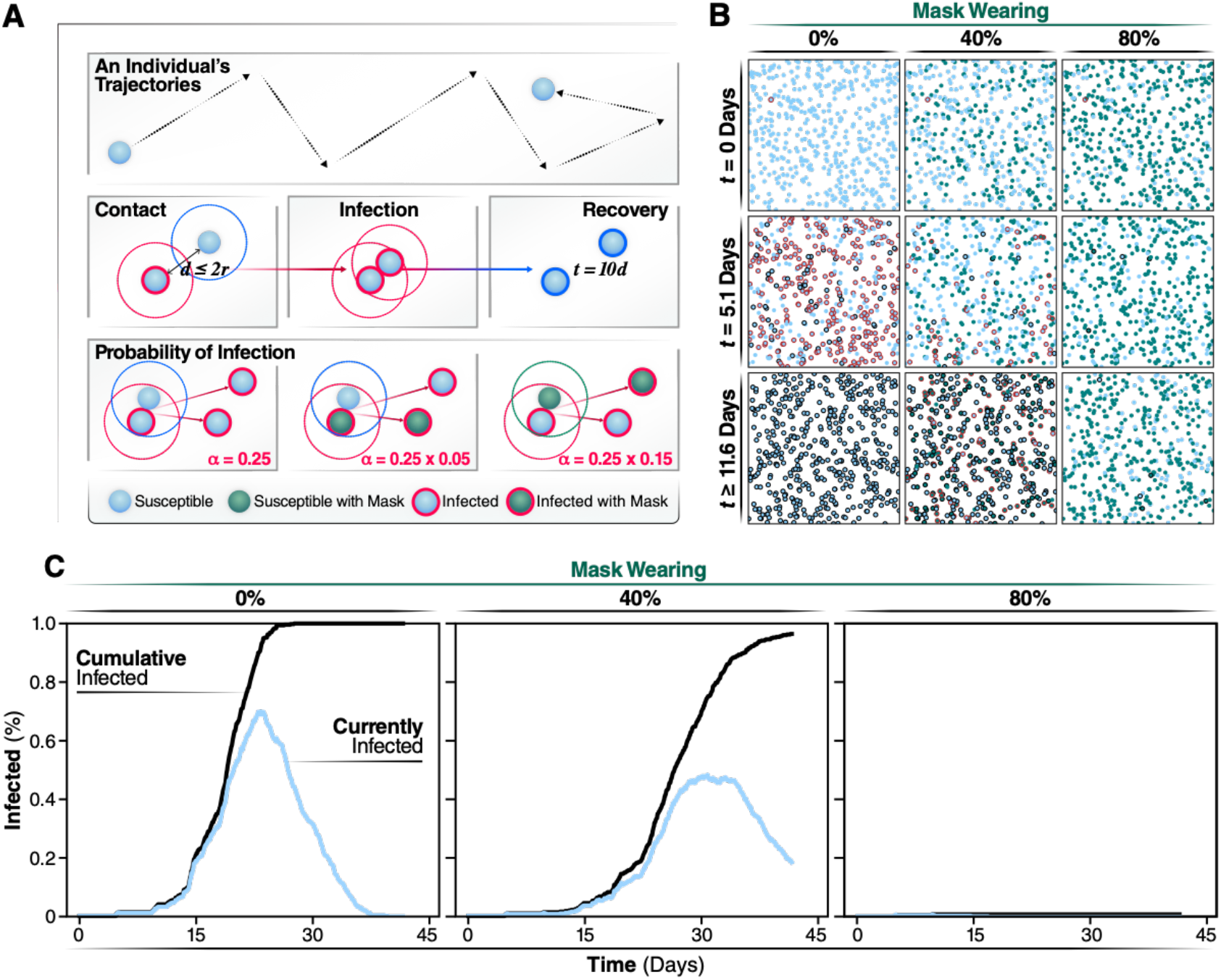
Individual states and rules of interactions. Using the same coloring system used in animated simulations, we show the different states an individual can be in during a simulation. Individuals keep the masked or not masked attribute assigned at initialization during the course of the simulation. **A**) Spread of infection is caused by interactions of overlapping trajectories between infected and susceptible individuals. If the distance between individuals, d, is less than the radius of the two individuals, r, then an interaction occurs. Interactions initiate the generation of a random number that determines if transmission occurs. The chance of infection is randomly generated, based on a gamma distribution with shape parameter, alpha, of 0.25. This probability is further modulated by which individuals in the interactions are wearing face masks. **B**) Agents are shown at their positions and states during snapshots of the simulation. Representative simulations of are shown of 0, 40, or 80% of the population wearing face masks. Snapshots are collected at days 0, 20, and 40 of the simulations. **C**) Progression of the outbreak, the trajectory of the number of new daily infected individuals (Currently infected) and cumulatively infected from a representative simulation are graphed in light blue and black, respectively.

To define the probabilities of infection, we used reported COVID-19 parameters^24^. The probability of transmission follows a Γ *(gamma)* distribution (see Methods) whose shape is described by a constant (α), estimated to be 0.25. Wearing masks reduces this probability (Fig. 1, probability of infection). To estimate the protective effect of masks, we used parameters determined for FDA-approved surgical masks, whose efficacy has been experimentally verified to inhibit virus transmission^10^. Based on previous studies (Leung et al.)^9^, we assumed that, if an infectious individual wears a mask, the effective probability of transmission is reduced by 95%. If a susceptible individual wears a mask, α is reduced by 85% (Fig. 1A). Our assumption that wearing masks is more effective to reduce transmission than to prevent getting infected is supported by experimental data^25^. However, if both infectious and susceptible individuals wear masks the probability of transmission is the product of these probabilities (0.0075) and, thus, sufficiently low such that transmission is effectively null. We calibrated our model by running simulations without any individuals wearing a face mask or practicing SD and considering that 50% of infected individuals are asymptomatic at the beginning of the simulation, with each new infection throughout the epidemic having a 50% probability of being asymptomatic (Movie 1). In this way, we determined the simulation parameters, such as velocity and density of individuals, to obtain a value of R_0_ = 2.5, consistent with what was reported early on as the infection rate of the epidemic in Wuhan^9^.

### Percentage of population wearing masks determines the daily infection incidence and cumulative number of cases

We started each simulation with one individual being infected and all others susceptible and assuming all subsequent infections have a 50% chance of being an asymptomatic. Non-infected and asymptomatic individuals circulate in the population without any restriction (i.e., they do not isolate themselves or become hospitalized) (see Methods). In contrast, symptomatic individuals no longer move after 12 hours of the symptom onset, simulating hospitalization or self-isolation. Thus, symptoms are assumed to manifest after the incubation time of 5.1 days (Fig. 2B). Using these assumptions and model calibration, we carried out a set of simulations in which we gradually increased the percentage of individuals in the population that wear masks. Individuals, who are assigned randomly to wear a mask at the beginning of the simulation, keep on the mask for the entire duration of the simulation. Increasing the fraction of the population wearing face masks has a highly significant effect on the spread of the virus (Fig. 1B, 1C and Movie 2). Mask wearing reduces the cumulative number of infected individuals at the end of the simulation (Fig. 1B). Strikingly, we observed a negative correlation between the percentage of the population wearing masks and the overall number of cases (Fig. 1B). The description of the dynamics of infection generated by this model is consistent with previous clinical studies^9,25^ and highlights the benefit of wearing masks (Fig. 1C, Movie 1).

**Figure 2.**
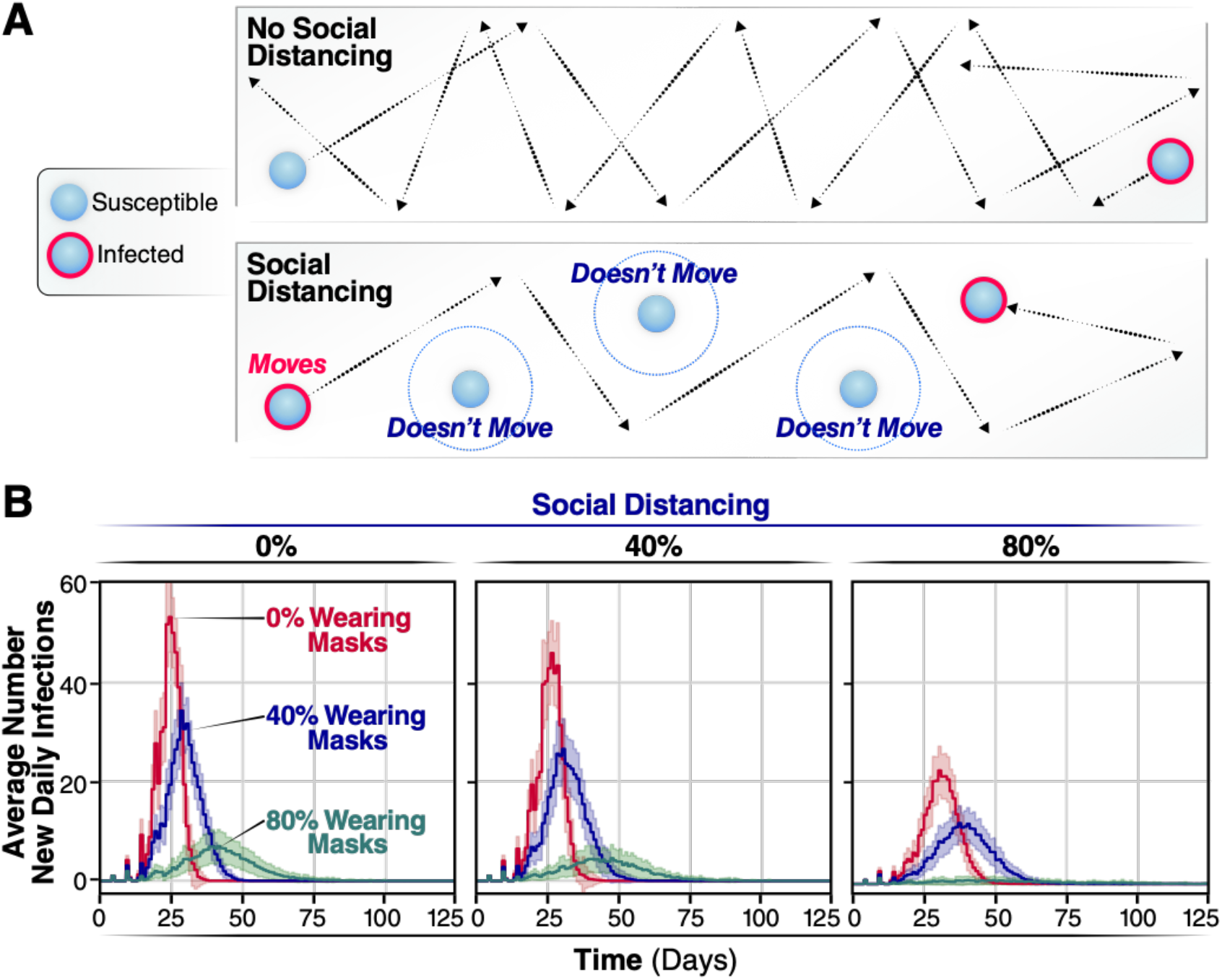
Average new infections per day when varying the population wearing masks and practicing social distancing. **A)** SD is implemented by assigning agents to not move along a trajectory during the simulation. This lack of movement reduces the number of trajectories overlaps between agents and subsequently reduces the number of transmission events. **B)** The number of new infections per day for 0, 40, or 80% of a population wearing face-masks are displayed by red, blue, or green trajectories, respectively, for 0% of the population practicing SD, for the 40% and for the 80%. Simulations were repeated 100 times for each condition, the curves and the highlighted regions around the curves represent the mean value +/-one standard deviation.

If the daily incidence surpasses the treatment capacity, it will overwhelm the healthcare system with detrimental consequences for medical care of infected individuals and increased mortality and morbidity. Thus, we examined the effects of face masks on the daily incidence of infection over time (Fig. 2B). If all individuals move freely and randomly interact with others (i.e., 0% SD), the rate of daily infection through the population depends on the percentage of individuals wearing masks. The average number of new daily infection varies considerably, according to the number of individuals wearing face masks.When 0% of individuals wear face masks (Fig. 2b, red line), the number of daily infections peaks sharply at day 27 with a maximum of 54.8 (51.2, 58.4 95% C.I.) infections per day for a population of 500 individuals. No additional infections are observed after day 39, when the entire population (100%) has been infected. Thus, without any intervention, the infection quickly reaches every individual through an epidemic characterized by a very sharp infection peak. Note that, in these original models, we assume all individuals recover after ∼11 days post-infection and cannot be re-infected. When 40% of all individuals wear masks, the number of individuals infected at any given day is reduced by approximately 30% (maximum of infected individuals 35.1 (32.6, 37.7 95% C.I.) Fig. 2A, blue line), both flattening the curve and extending the duration of the outbreak by more than 10 days, with a peak maximum at day 32. By day 52, an average of 472 individuals has been infected, and none of the individuals is infectious or exposed any longer. Even more significantly, if 80% of the population wear masks, we observed a significant flattening of the curve, with a substantial reduction in the maximum number of infected individuals per day, 5.9 (4.6, 7.3 95% C.I.) (Fig. 2A, green line), and the number of new infected individuals reached zero by day 57.8 ± 35.0. Thus, the shape of the outbreak changes from a curve characterized by a sharp peak when no intervention is considered to a broader peak when 80% of the individuals wear masks. By increasing the percentage of the individuals wearing masks, the number of newly infected individuals per day substantially decreases, which will reduce mortality and morbidity. Moreover, since the use of masks eliminates the sharp peak that characterizes SARS CoV2 epidemics, the overall impact of the outbreak on the health system is alleviated. These results highlight the importance of widespread mask wearing as an effective intervention that can be implemented as soon as the first cases are reported.

### Effect of social distancing on viral infection spread

Next, we evaluated the effect of SD in shaping the spread of infection. Practicing SD does not affect the probability of infection conditional on an encounter but reduces the chances of encounters leading to transmission. Thus, SD was introduced into the model by limiting the proportion of individuals in the population that move freely in the field. This simulates a scenario in which a given proportion of individuals in the population quarantine or shelter-in-place, thereby reducing the probability of contacts and transmission (Fig. 2A). As reported, if a percentage of individuals practice SD, even without any other non-pharmacological intervention, the number of infections and daily infection rate are reduced (compare Fig. 2B, SD 0, 40, and 80%, red line). However, if no one wears masks but 40% of individuals practice SD, we observed a very small effect in the shape of the infection curve (Fig. 2B, the peak maximum decreases from 51.0 (48.3, 53.6 95% C.I.) to 30.1 (27.5, 32.7 95% C.I.) (). When 80% of the population practices SD, a more significant reduction in the number of new daily infections was observed (Fig. 2B, 80%). Notably, the shape of the new daily infection curve broadens considerably upon increasing the percentage of individuals wearing face masks (Fig. 2B, compare blue and green curves with red curve). For instance, if 80% of the population wears masks with 40% SD, the peak maximum decreases to one-tenth (from 51.0 (48.3, 53.6 95% C.I.) to 5.7 (4.5, 6.7 95% C.I.)) and is slightly delayed (Fig. 2B). At 80% SD, the peak of new daily infections is no longer observed, and the number of new infected individuals per day averages 1.6 (1.4, 1.9 95% C.I.) (Fig. 2B). Thus, the effects of individuals wearing face masks and practicing SD are synergistic, with the most pronounced effects occurring when 60% or 80% of individuals wear masks.

It is possible that our results may vary depending on the way social distancing is implemented in the model. To test this possibility, we carried out additional simulations in which modulate the amounts of social distancing by completely removing infected agent practicing SD from the simulation, and only allowed infected agents to be symptomatically infected (see SM 6). The results from these simulations were similar to those described above, confirming that around ∼60-80% of the population is required to practice SD to effectively slow the rate of infection, even when SD reduce the probability of transmission to zero. Our results also show that, if infection is always symptomatic, the number of infections drops significantly, which is consistent with the spread driven by asymptomatic individuals.

### Effectiveness of combining mask wearing and social distancing to control infection in populations with high proportions of pre-symptomatic and asymptomatic cases

Asymptomatic or pre-symptomatic SARS-CoV-2 infection is emerging as possibly the most common clinical manifestation of COVID-19^26-30^. This finding could only be revealed once mass testing campaigns were performed, regardless of symptoms (e.g., universal testing campaigns). One of the earliest studies documenting clinical manifestations in a testing campaign (which still focused testing mainly on symptomatic individuals) was on the *Diamond Princess* cruise ship, where the rate of asymptomatic infection was 18%^31,32^. In a mass testing campaign in Iceland, where testing was offered to a segment of the general population (regardless of symptoms), 43% of individuals were asymptomatic at the time of testing^33^. Thus, the actual percentage of pre-symptomatic and asymptomatic cases is currently unknown, but it is clear that a large number of SARS CoV2 new infections derive from undetected infections. Controlling the outbreaks by isolation or even by increased population testing is a big challenge and may be difficult to implement. We thus determined the efficacy of mask wearing and SD in the context of different proportions of asymptomatic incidence. We assume that asymptomatic individuals are less infectious than symptomatic individuals: asymptomatic individuals may have lower viral loads and reduced coughing, sneezing, and nasal secretions, all of which may facilitate transmission^34-36^. The model considers that symptomatic infected individuals isolated themselves 12 hours after the onset of symptoms, because they get hospitalized or self-isolated. In contrast, asymptomatic individuals remain infectious, circulate and transmit for a period of 7 days (Fig. 3A). Because some individuals can be pre-symptomatic (i.e., symptoms emerge later after the initial incubation time (Fig. 2B, *t= 5*.*1* days)), we modelled infectivity of asymptomatic/pre-symptomatic as linearly declining until the individual is no longer infectious, after 7 days^37^. Using these assumptions, we performed a sensitivity analysis by carrying out simulations, varying the percent of asymptomatic/pre-symptomatic cases to determine the efficiency of non-pharmacological interventions in populations with 25, 50, and 75% pre-symptomatic and asymptomatic infected individuals (Fig. 3).

**Figure 3.**
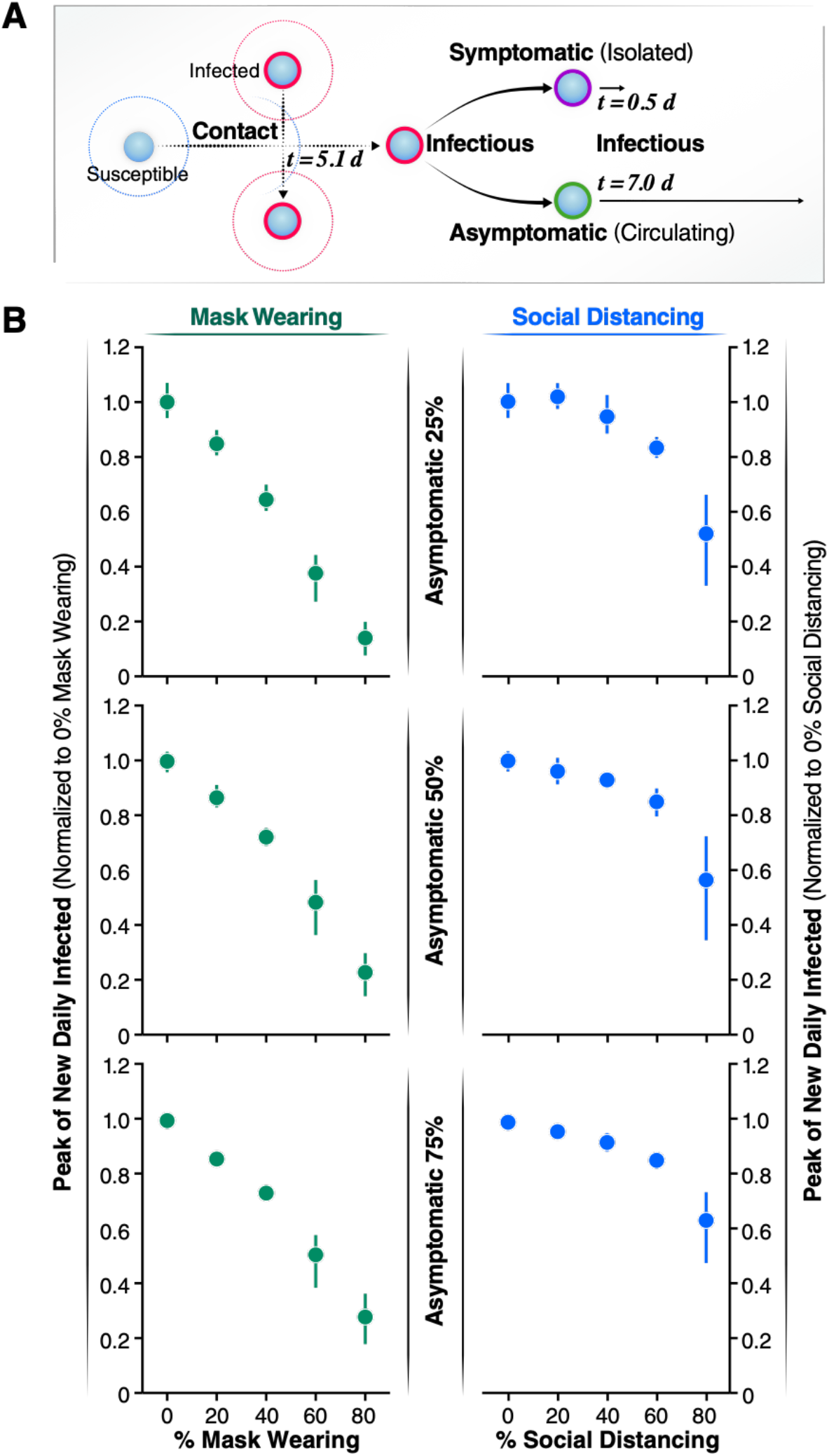
Effects of the asymptomatic population on the infected peak number. **A**) Schematic representation of the asymptomatic transmission during the simulations. After 5.1 days of coming into contact with an infected individual, infectious agents can develop symptoms and after 12 hours will be isolated. On the contrary, asymptomatic individuals don’t develop symptoms of the disease so they are not isolated but keep circulating thus contributing to spread the infection for 7 more days after the initial contact with an infectious agent. **B**) The normalized peak of new daily infected is represented as a function of the percentage of the population wearing mask (left panels, green solid circles) and as a function of increasing the percentage of asymptomatic individuals (25% top left panel, 50% central left panel, 75% bottom left panel). The normalized peak of new daily infected is represented as a function of the percentage of the population practicing social distance (right panels, blue solid circles) and as a function of increasing the percentage of asymptomatic individuals (25% top right panel, 50% central right panel, 75% bottom right panel). Error bars represent 95% confidence intervals calculated.

Our simulations demonstrate that the total number of infected individuals increases linearly with the increase of the percentage of asymptomatic individuals in the population (Fig. 3B). To compare each condition, we normalized the peak of new daily infected individuals to conditions in which no one practiced SD or wore masks. Strikingly, increasing the percentage of individuals wearing masks linearly reduces the normalized peak number of infected individuals per day (Fig. 3B). For instance, when 40% of the individuals wear face masks, the peak number of infected decreases similarly independently of the percent of asymptomatic cases considered (to 0.6–0.7, Fig. 3B). Furthermore, the linear relationship between the decrease in peak number of newly infected individuals per day and increase in percent individuals wearing masks is independent of the proportion of asymptomatic individuals in the population (Fig. 3B). This finding indicates that, when the fraction of asymptomatic cases is high, as in the case for COVID-19, wearing face masks is as effective to reduce the peak number of infected as when a low percentage of individuals are asymptomatically infected.

In contrast, SD was only effective in populations with high incidence of asymptomatic infections when a very high fraction of the population practice SD (more than 60%) (Fig. 3B). Furthermore, the low efficacy of SD as a containment strategy is more pronounced when the proportion of asymptomatic individuals increase (compare 25, 50, and 75%). Of note, if the number of individuals wearing face masks is high, increasing the number of individuals practicing SD has negligible effects on the daily number of infected individuals (see supplementary materials). One noteworthy observation from our model is that, at high rates of SD (e.g. 80 %), there is more variability.

Our analysis uncovers a linear relationship between the fraction of a population wearing masks and the reduction in infection rate. In contrast, we find SD requires a high fraction of compliance to be effective. These findings indicate that having a high percentage of individuals wearing face masks is more beneficial in preventing virus spread and reducing the peak number of infected individuals than having a high percent of people practicing SD. Notably, the benefit of wearing masks is not affected if the percent of pre-symptomatic and asymptomatic increases.

### Interplay of face masks and social distancing for controlling infection spread and protecting from COVD-19

Next, we determined the average cumulative incidence as a function of the percentage of individuals wearing masks and practicing SD (standard deviation described in Fig. SM3-A). When neither SD nor masks were used our simulations indicate that up to 99% of the population will end up infected, leading to unacceptable levels of mortality and morbidity. When the proportion of the population wearing masks was increased, no significant effect on the total number of infected people was noted until more than 40% of the population wore masks. However, if 80% of people use masks, the average cumulative incidence of infection decreased to around 35%. Therefore, wearing face masks alone would greatly limit the spread of the virus. In contrast, SD alone does not have a significant effect. Increasing SD compliance to 80% only reduced total infections by 8%, and 87% of the population was eventually infected. Importantly, the combination of wearing masks and practicing SD by a high proportion of the population dramatically reduces the total number of infected individuals to 10% of the population (Fig. 4A; and Fig. SM2-C).

**Figure 4.**
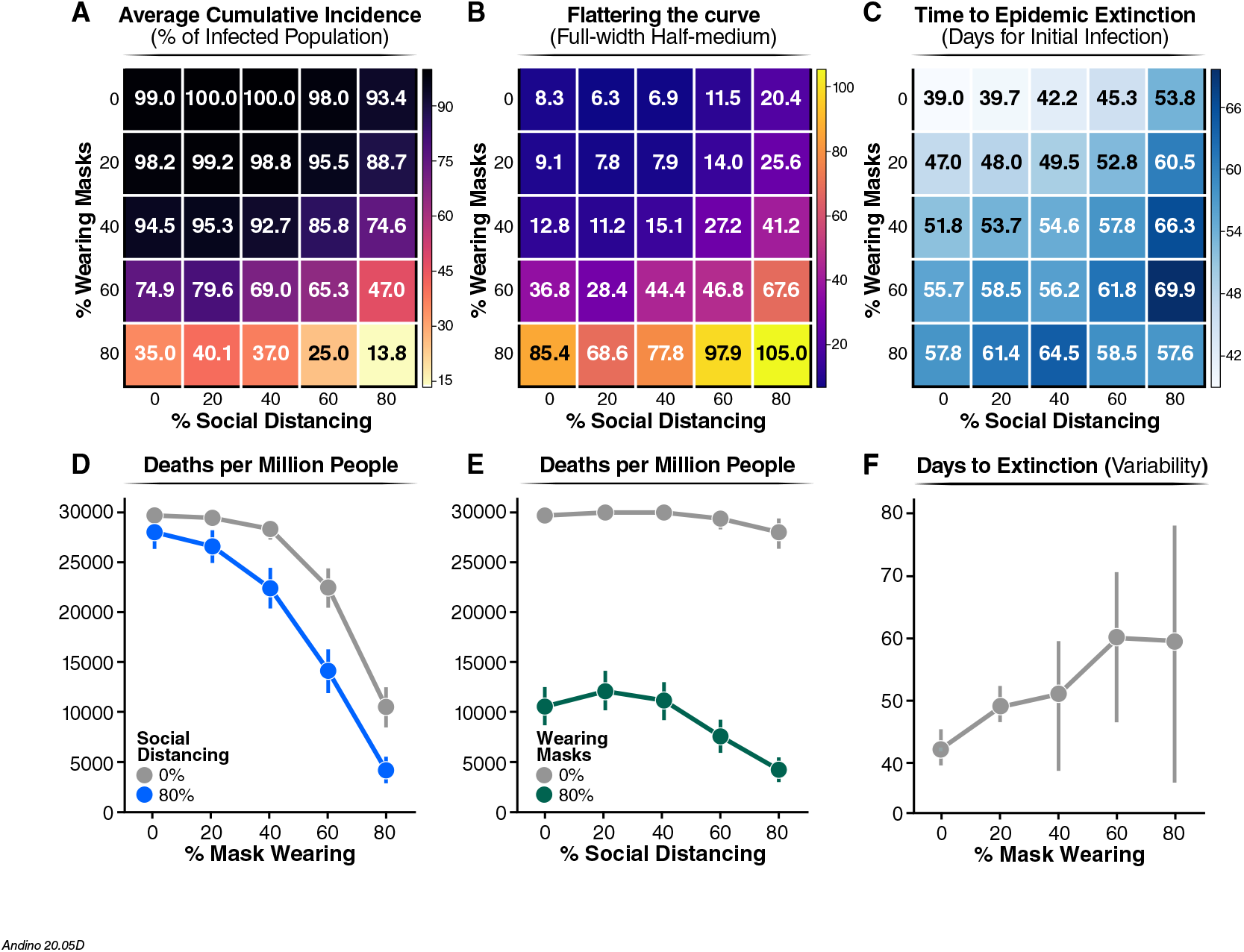
Summary values from 2500 simulations of varying percentages of population wearing masks or social distancing. **A**) Proportion of infected population. The average cumulative incidence is represented as a function of the population practicing SD or wearing a mask, which are given by the x and y axis, respectively. **B**) The shape of the daily new infection curve. Full-width half-maximum (FWHM), denoting the number of days between the first day and last day of cases that have half the peak number of infected individuals. **C**) Extinction rate of the epidemic estimated as the average number of days for which the simulation reports no new infected individuals. In the figure, the numbers represent the mean value calculated over 100 of simulations carried out for each condition. For clarity, we reported the standard deviation of the mean for each value reported in this figure in Fig. SM5. **D**) and **E**) Number of death people.Extrapolated for one million individuals from the cumulative incidence of infections estimated from our model at a mortality rate of 3%. **F)** Number of days from the start of infection spread to the last infected agent recovering. Error bars represent 95% confidence intervals calculated for **D**), **E**) and **F**).

To gain additional insights into the characteristics of the epidemic in response to these mitigation strategies, we studied the shape of the epidemic curve by calculating the peak and full-width half-maximum (FWHM) of the peak of the new daily infections (Methods). While the peak reports on the maximum number of daily infections at the height of the epidemic, the FWHM is a simple way to represent the duration of the characteristic peak of new daily infections. Indeed, FWHM reports on the extinction time of the infection within the population (number of days until no individual is still infected) (Fig. 4A, and Fig. SM3-B standard deviation). A larger FWHM corresponds to a flatter but longer epidemic curve. A flatter epidemic curve enables better management of the epidemic, as the healthcare system is not overwhelmed by the number of cases at a given time. Initially, the FWHM was calculated assuming no individual practice SD. FWHM was ∼8.3 days if none of the individuals wears masks, ∼12.8 if 40% of individuals wear masks, and but increases dramatically to 85.4 when 80% of the population wears masks (Fig. 2B). Our data indicate that wearing masks has a more profound effect than SD on flattening the epidemic curve. For instance, when 80% of the population wears masks, the epidemic curve is eight times flatter than without any non-pharmacological intervention (Fig. 4B). In contrast, if 80% of the population practices SD, the flattening of the curve is less than threefold. The most dramatic effect on flattening of the curve is observed when wearing masks is combined with SD; for instance, if 80% of the population wears masks and 80% practices SD, the curve is flattened over 10-fold, compared to no intervention.

We also find that the reduction in the number of infected individuals per day correlates with a lengthening of the outbreak. Broadening of the peak affects the extinction time of the infection (Fig. 4C).For instance, when the percentage of individuals wearing face masks rises from 0 to 80% of the population, the extinction time increases from ∼39.0 to ∼58 days (Fig. 4C). Similarly, as a higher proportion of individuals practice SD, the time to extinction of the infection also increases (Fig. 4C, ∼39 to ∼54 days for 0% to 80% of SD). Importantly, even though the time to epidemic extinction is extended, the total number of infected individuals dramatically decreases (Fig. 4A). If a higher proportion of the population (80%) wears face masks and practices SD, the time to epidemic extinction is reduced (Fig. 4C) because the total number of infected individuals is dramatically reduced (Fig. 4A). Thus, stochastic effects dominate, and this can be appreciated by the wider error bars (Fig. 4F).

To relate the impact of these interventions to their societal impact, we determined the number of deaths per million after each mitigation strategy, assuming a mortality rate of ∼3%^38^. This analysis illustrates the heavy cost of lives of the virus, but also demonstrates that a high level of mask wearing compliance is the most effective non-pharmacological approach to protect human lives, particularly when combined with even moderate SD measures (Fig. 4D). In contrast, SD, without masks wearing is not effective to reduce mortality (Fig. 4E). Finally, our simulations predict that increasing the proportion of the population wearing masks will increase the time to outbreak extinction (from ∼40 to ∼60 days) (Fig. 4F).Together with the broadening of the peak (Fig. 4C), this shows an effective flattening of the curve. Importantly, with 80% mask wearing, we observed an increase in the statistical distribution from the average time to extinction (Fig. 4F, see 95% confidence intervals). Thus, a generally low disease incidence triggers stochastic events leading to extinction of the infection. Our simulations represent real outbreak scenarios and reveal that as the outbreak approaches its extinction there is an increase in the uncertainty of whether or not the infection has been completed eliminated, which argue to be prudent before society reopening can be done safety.

## Discussion

Here we use realistic simulations rooted in experimentally measured parameters of SARS-Cov2 spread, contagion mode and mortality, to evaluate two available NPIs that reduce the spread of a respiratory infection, such as COVID-19. In our simulation, we assumed proper use of FDA-approved face masks. We showed that a high degree of compliance in the use of masks, regardless of whether the wearer displays symptoms, slows the spread of infection. Face masks substantially reduce the transmission of respiratory droplets and aerosols containing viral particles^9-11^. Increasing the fraction of the population wearing face masks reduces the number of new infected individuals per day and flattened the curve of total individuals infected (Fig. 2A and 4A). These two effects should reduce mortality and morbidity, alleviate the current stress on healthcare systems, and enable a more effective management of severe cases. However, solely wearing masks cannot entirely prevent an outbreak from occurring. It cannot by itself extinguish the virus, since as long as a small fraction of the population is non-compliant, the virus can persist in the population. Our models show that combining proper use of masks with practices such as SD, indisputably decreases the number of new infected individuals per day (Fig. 2). The asymmetry between the effectiveness of SD and mask wearing is of particular interest. In a classical, ODE-based epidemiological model, mask wearing and SD would both affect the transmission rate. In the absence of SD, imagining a 100% effectiveness of mask, the reduction in transmission due to mask wearing would be the same as the decrease due to SD in the absence of mask wearing. In fact, both controls would be, effectively, modeled as a removal of people from the population. On the other hand, our simulations show that mask wearing has a stronger effect than SD on the disease incidence. This is due to the way we model SD, that is, as a decrease in mobility. A better approximation of our model is to consider an epidemic dynamic on a scale-free network, where homogeneous mixing is relaxed and SD is modeled as a change in the degree distribution of the network^39^. In addition, our model was demonstrated to be scalable and able to describe realistic situations such as interactions between 5000 agents (see supplementary materials).

Our analysis provides guidance for policies to protect the population from COVID-19. Optimizing the use of masks with SD practices effectively limits the virus spread and reduces several parameters in the epidemic, including cumulative incidence, shape of the peak, and the extinction rate (Fig. 3). In particular, we observed that wearing masks is more effective than SD. Even in a population with a high number of asymptomatic infections, increasing the use of masks up to 80% results in a significant reduction in infection (Fig. 4A). Meanwhile, even 80% of individuals practicing SD has only a marginal effect (Fig. 4D). This result can be understood in terms of contact rate, since we assume that asymptomatic infectious individuals have higher mobility than symptomatic infectious ones. If the vast majority of the population is asymptomatic, then high compliance with face mask use is a key factor for curbing the epidemic. Moreover, we believe that our methods are general and they may inform policies against other respiratory infections like influenza.

Our simulations also provide insights into how enforcing different mitigation practices affects the length of the epidemic. Assuming a homogenous population, the trajectory of epidemic extinction lasts 50–60 days, when 80% of the population either wears masks or practices SD (Fig. 4C). However, when 80% of the population is wearing masks and 0% of the population is practicing SD, the cumulative incidence is reduced three times (∼35%), and the peak is very broad (FWHM of ∼ 85 days). In contrast, when the population solely practices SD (80%), the majority of the population (93%) will end up infected, and the peak of daily infected individuals will be sharper (FWHM of ∼ 20 days).

Our model indicates that the synergistic utilization of face mask wearing and social distancing practice is most effective in controlling SARS-CoV-2 spread. We observed that wearing masks in combination with some degree of SD relaxes the need for a complete lockdown, leading to a potential suggestion for an intervention policy based on a mix of the two measures. The effectiveness of mask wearing to control virus spread is not reduced if a large fraction of the population is asymptomatic. This suggests that, in the absence of universal testing, widespread use of face masks is necessary and sufficient to prevent a large outbreak. Our results are supported by the real data of Korea^33^ and Taiwan^34^, where an early mandate to requiring face mask usage, in combination with SD, severely limited the spread of the virus. While more work is necessary to specifically assess the impact of other variables shaping COVID-19 outbreaks, such as increased mobility, age stratification, testing a fraction of the population, our study can accurately inform strategies to reduce the spread of the virus. In particular, our results may be highly relevant toward informing specific realistic situations, such as the spread of the disease in a confined space, where effective SD may not be easily achievable (e.g., schools, essential businesses, correctional facilities, public transportation, hospitals). These strategies, if effectively implemented, will save countless lives from the SARS-Cov2 infection. According to our model, if the United States (330 million people) does not implement any NPIs, then ∼627,000 people are expected to die. Contrastingly, if 80% of the population wear face masks, that number would significantly shrink to about 250,000. If both face masks and SD were practiced by 80% of the population from the start of a pandemic, the mortality rate decreases to 65,600 people.

## Materials and Methods

We developed and used our python codes using the NumPy library version 1.15.4^40^ to carry out ABM and describe the dynamic evolution of a SEAIR system affected by COVID-19 disease, in which each individual can be in a susceptible (S), exposed (E), asymptomatic infected (A), symptomatic infected (I), or recovered (R) status. In addition, each individual can wear a face mask (M) or can practice social distancing (SD), where wearing face masks or practicing SD are independent binary values of an individual. In the simulations, each individual was represented by a position in a 2-D lattice of 21×21 dimension with periodic boundary conditions. Individual initial positions are assigned randomly by NumPy’s random module, and the simulations start with all susceptible individuals and only one exposed. During the simulation, each individual move along a randomly oriented trajectory at a constant velocity, moves the same distance between each time step of the simulation, and interacts with individuals whose position is within a fixed diameter of another individual. The data from each individual are saved as vectors of attributes, including position, velocity, state (S, E, A, I, R, M, SD), and number of individuals they come into contact with at every simulation time step. A time step corresponds to 1 hour. We implemented the state of M by defining sub-routine during the interaction between two individuals where the probability of infection was reduced by an amount corresponding to which of the two individuals is wearing a mask. For the SD state, we assigned the individual to be stationary and not follow along a randomly assigned trajectory.

We refer to Ferguson et al. for defining the rules governing the interactions among individuals^24^. For the interaction between symptomatic infected and susceptible individuals, the infection probability was randomly sampled from a gamma distribution with a mean of 1 and a shape of 2.5, whereas for that between asymptomatic infected and susceptible individuals, the infection probability is reduced by 33 %. We assume this reduction of infectivity for the asymptomatic, based on the absence of transmission-aiding symptoms, such as coughing, sneezing, and a runny nose^24^. The time between a susceptible individual being exposed to being in the infectious infected state is 5.1 days^24^. Pre-symptomatic agents are defined as infected agents during the incubation period. Symptomatic infected individuals are infectious for 12 hours before self-isolating. After this time, we assume that these individuals no longer infect those around them because they are hospitalized or self-isolating. Asymptomatic infected individuals are infectious for 7 days, and the infectivity linearly declines until the individual is no longer infectious on day 7^37^. If an infected, a susceptible, or both individuals are wearing a mask their probability of another individual being infected is reduced to 5, 15, or 0%, respectively, from the original gamma distribution.

First, we carried out a simulation with 500 individuals with 0% of individuals wearing face masks and 0% practicing SD to optimize parameters as velocities and individual density to obtain a basic reproduction number, R_0_, equal to 2.5, based on reported values. The basic reproduction number was computed from the mean number of symptomatic cases, resulting from a single symptomatic individual. Then, we carried out four more sets of simulations in which we increased the percentage of individuals wearing masks by the 20, 40, 60, and 80%. Finally, for each of these simulation sets we carried out four more sets in which we increased the percentage of individuals practicing SD by the 20, 40, 60, and 80%. Simulations are stochastic, and we simulated each condition 100 times to increase the statistical power of the analysis. In total, we simulated 25 different conditions for a total of 2500 simulations, 100 for each condition. In all these simulations, the probability of a new infection being asymptomatic was 50%. Simulations were run until there were no individuals either infected or exposed. Summary values were computed by methods in the NumPy module. Error bars were calculated by the standard deviation method in NumPy and for figure 3 and 4 the error bars were calculated by 95% confidence interval. The full-width half-maximum (FWHM) was calculated for each simulation from the newly infected per day data by taking the maximum and using it to find the days that intersect the value of half the maximum value. The first and last days of this intersection were then used to calculate the number of days of the FWHM. From the set of FWHM for each simulation, an average and standard deviation were calculated using the NumPy module. FWHM error bars are large due to a large variation in how flat the curves are. Graphs were created by using the Matplotlib^41^ and Seaborn^42^ modules in python.

We followed the same protocol described above to perform simulations, varying the probability of a new infection being asymptomatic from 25 to 75%. Finally, to monitor the reproducibility of our results with a larger population and contrast with real data, we ran four sets of 30 simulations with 5000 individuals in the combinations of 0 or 80% SD and 0 or 80% wearing masks. All 5000 individual simulations were done with 50% asymptomatic infection rates.

As the position, trajectory, and state of each individual in the simulation are explicitly known, using the matplotlib library’s animation package, we converted each time step of the simulation to an image in a movie, representing 1 day as a second spanning the length of the simulation.

For each simulation, we saved in a .csv in tidy data format the summary values of the simulations, which we used for the analysis. The simulation was written in python and all scripts are available on GitHub at https://github.com/adamcatching/SARS_SEIR_Simulation, simulations were generated using script mask_single_sim.py.

## Data Availability

All the data is available at the link below

https://www.adamcatching.com/movie_2.mp4

https://www.adamcatching.com/movie_1.mp4

## Acknowledgments

We acknowledge the use of the UCSF Computing Cluster Wynton (https://wynton.ucsf.edu/hpc/index.html). We thank Judith Frydman, Alma Andino-Frydman, and Patrick Dolan for their suggestions on the manuscript.

## Author contributions

A. C., S. C., and M. T. Y. conceived the study. A. C. and S. C. designed the simulations. A. C. wrote and executed the simulations. A. C., S. C., and S. B. analyzed the data. S. B. and R. A. provided guidance and support. All authors wrote the manuscript.

