## Supplemental Materials for "Examining face-mask usage as an effective strategy to control COVID-19 spread"

[§] authors contributed equally to the manuscript

**Legend for the movies**

**Movie 1. Representative simulation with 0%, 40%, and 80% agents wearing masks and 0% practicing social distancing.**
Representative movies of the set of 100 simulation carried out with 0%, 40%, and 80% of population wearing masks. The dots are colored according to their status: *light blue* is susceptible, *red ring* is exposed and infected, *teal* is susceptible with mask. Simulations are carried out with 500 agents and with 50% of asymptomatic infected population.

**Movie 2. Representative simulation with 0%, 40%, and 80% agents wearing masks and 80% practicing social distancing.**
Representative movie of the set of 100 simulation carried out with 0%, 40%, and 80% of population wearing masks. The dots are colored according to their status: *light blue* is susceptible, *red ring* is exposed and infected, *teal* is susceptible with mask. Simulations are carried out with 500 agents and with 50% of asymptomatic infected population.

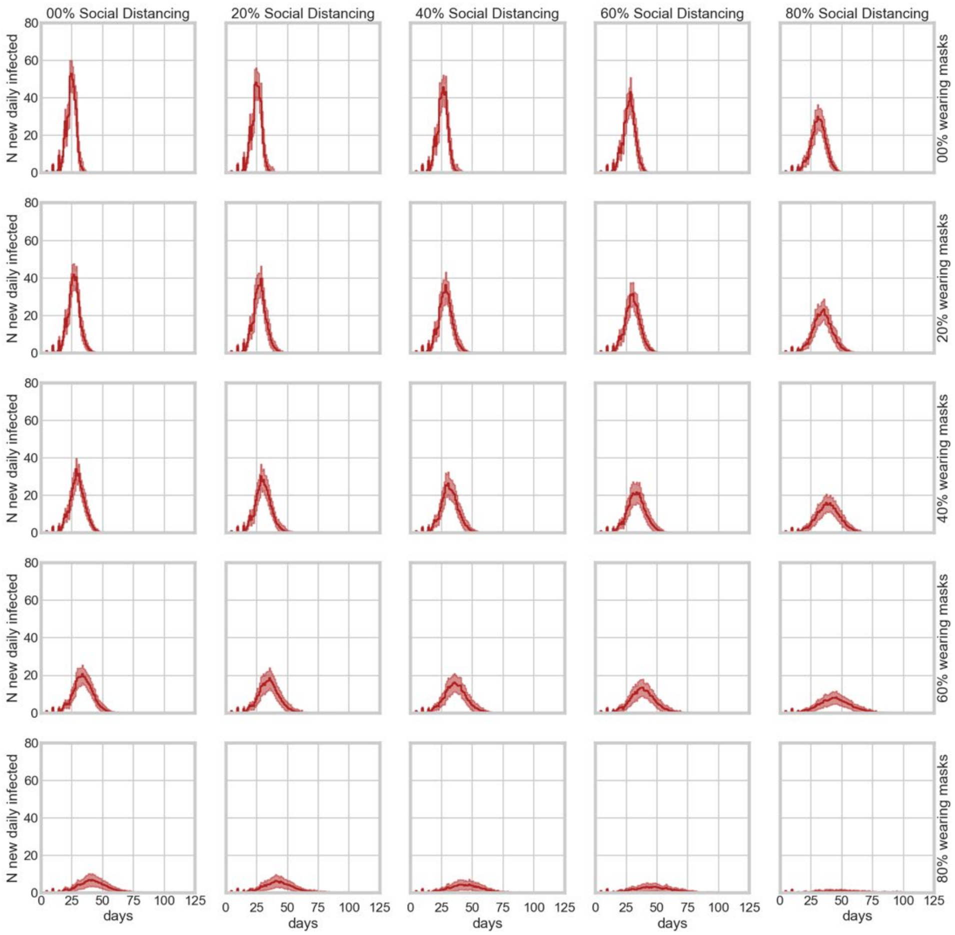


**Fig. SM1. Number of infected agents with 50% of asymptomatic infected population.**
Number of infected agents as a function of time for all combinations of different percentage of the agent population practicing social distancing (x-axis) and wearing masks (y-axis). 100 simulations are represented for each condition and each simulation is displayed as a low-opacity trajectory over time.

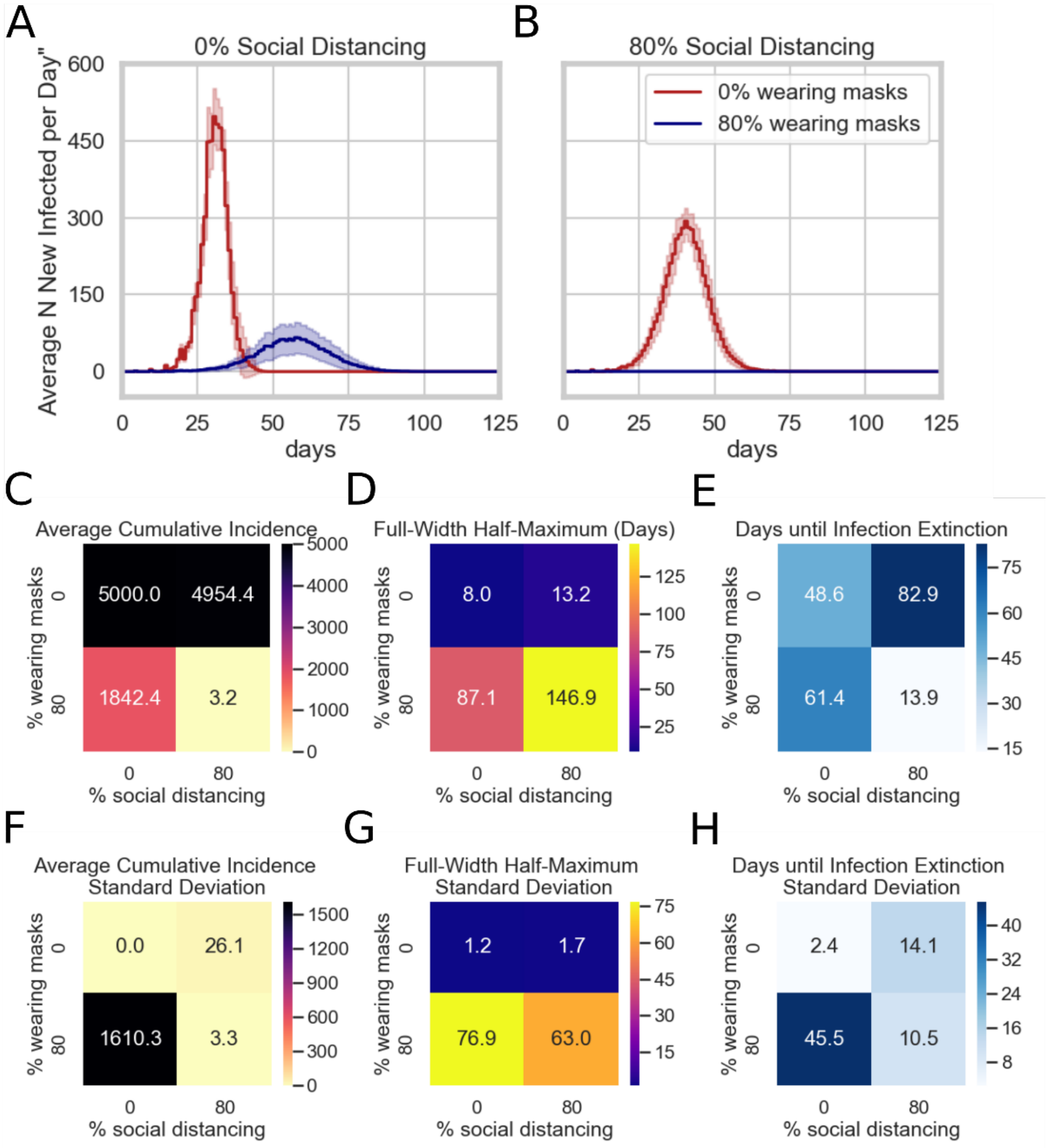


**Fig. SM2. Dynamics of the epidemic with simulations of 5000 agents**

Simulations were run with 500 agents. **A**) and (**B**) report the number of new infected agents per day calculated for either 0 % (red line) or 80 % (blue line) of the population wearing masks and for either (**A**) 0 % practicing social distancing or (**B**) 80 % practicing social distancing. **C**) The average cumulative incidence is represented as a function of the population practicing social distancing or wearing a mask, which are given by the x and y axis, respectfully. **D**) Full-width half-maximum (FWHM), denoting the average number of days between the first day and last day of cases that have half the peak number of infected agents. **E**) Extinction rate of the infection estimated as the average number of days for which the simulation reports no new infected individuals. Data for each condition was simulated 30 times and the highlighted regions around lines in figures **A**) and **B**) represent the mean value +/- one standard deviation. **F**), **G**), and **H**) represent the standard deviation of the mean reported in **C**), **D**), and **E**)

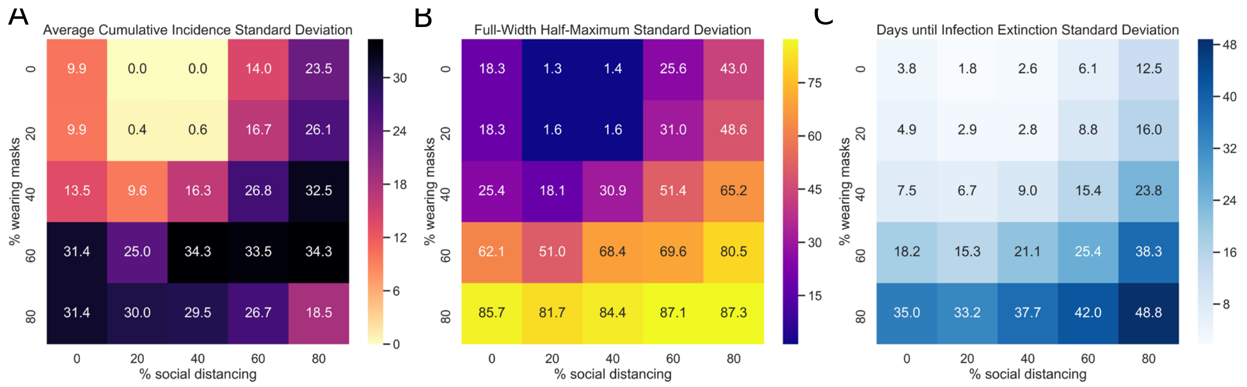


**Fig SM 3. Standard deviation of summary values from 500 agent simulations varying percentages of population wearing masks or social distancing.** Each condition was simulated 100 times and standard deviation of the mean values reported in Figure 2 is represented. **A**) The standard deviation of the average cumulative incidence is represented as a function of the population practicing social distancing or wearing a mask, which are given by the x and y axis, respectfully. **B**) The standard deviation of the Full-width half-maximum (FWHM), denoting the standard deviation number of days between the first day and last day of cases that have half the peak number of infected agents. **C**) Standard deviation of the extinction rate of the infection estimated as the standard deviation number of days for which the simulation reports no new infected individuals.

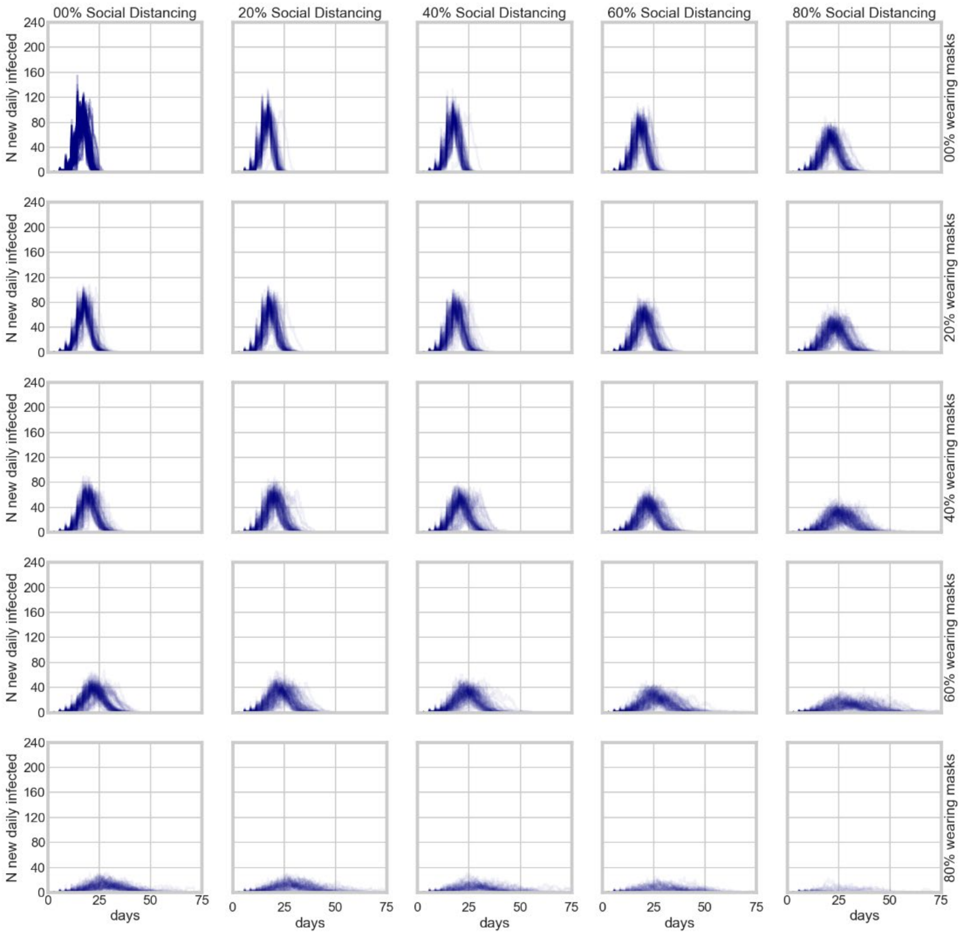


**Fig. SM4. Number of infected agents with 25% of asymptomatic infected population.**
Number of infected agents as a function of time for all combinations of different percentage of the agent population practicing social distancing (x-axis) and wearing masks (y-axis). 100 simulations are represented for each condition and each simulation is displayed as a low-opacity trajectory over time.

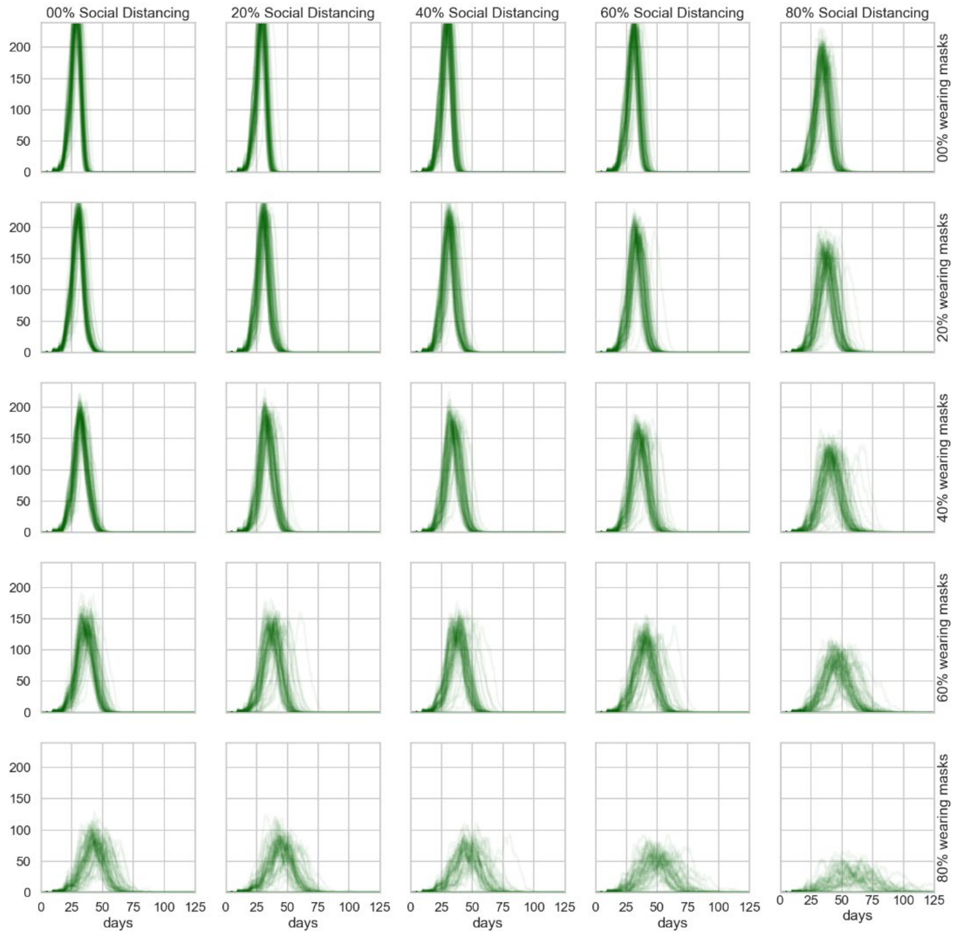


**Fig. SM5. Number of infected agents with 75% of asymptomatic infected population.**
Number of infected agents as a function of time for all combinations of different percentage of the agent population practicing social distancing (x-axis) and wearing masks (y-axis). 100 simulations are represented for each condition and each simulation is displayed as a low-opacity trajectory over time.

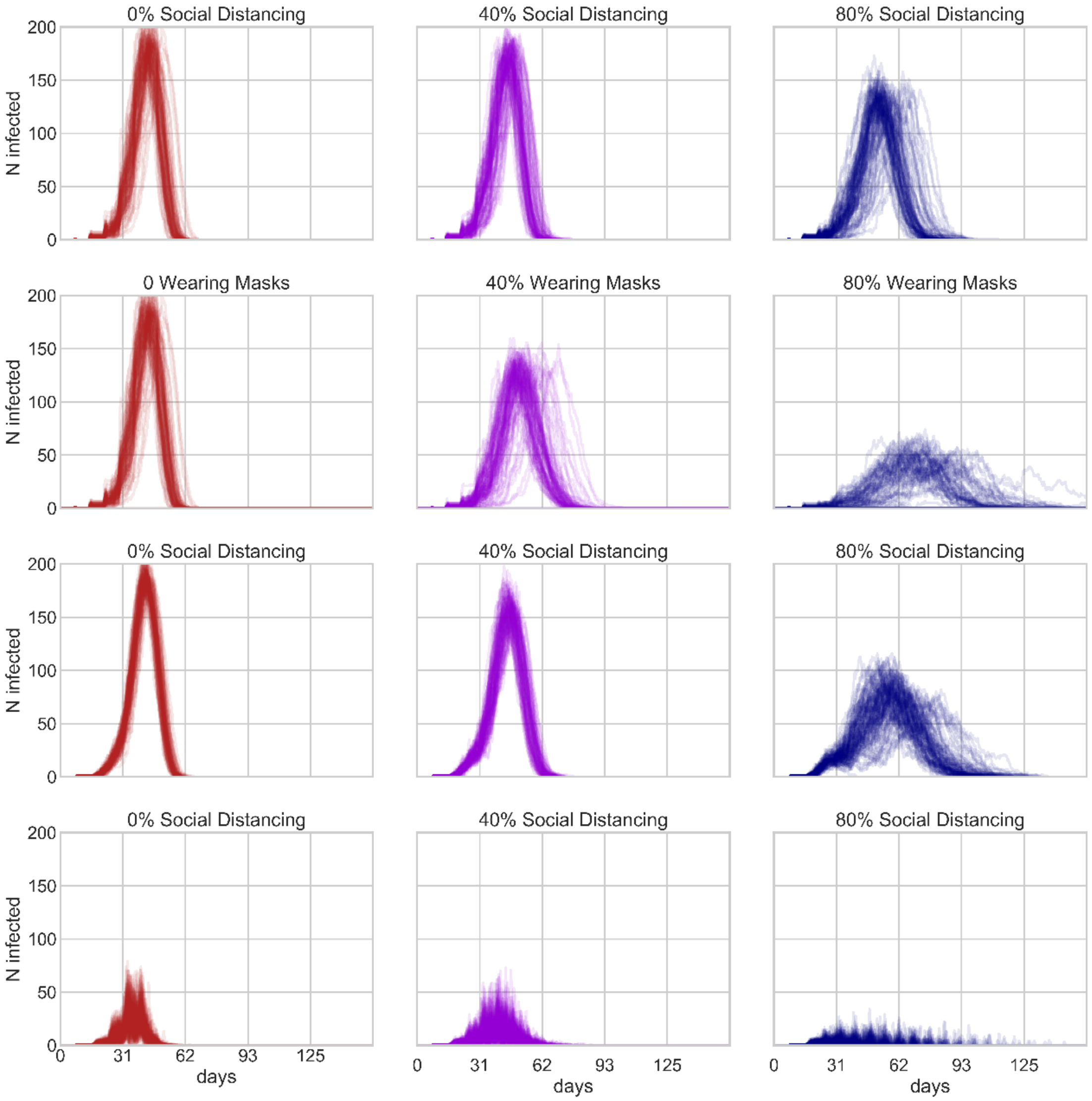


**Fig. SM6. Impact of social distancing hypothesis**

Measure of the impact of social distancing hypothesis on the number of infected agents as a function of time. 100 simulations are represented for each condition and each simulation is displayed as a low-opacity trajectory over time. The first row reports the baseline simulation with SD implemented as described in the main text and no mask wearing, for various percentage of SD. The second row reports the simulations for various fraction of mask wearing and no SD. The third row reports the case of removal from the simulation of individuals practicing SD, no mask wearing. The fourth row uses the same simulation method as the third row, except infected agents can only become symptomatically infected.
